# The timeline of brain aging in major depression: A prospective study from before first-onset to established illness

**DOI:** 10.64898/2026.08.29.26361710

**Authors:** Maximilian Konowski, Anna Kraus, Janik Goltermann, Jan Ernsting, Keyvan Mahjoory, Lukas Fisch, Jennifer Spanagel, Sarah Wellms, Dunya Bedir, Luisa Altegoer, Tiana Borgers, Kira Flinkenflügel, Sabrina Teckentrup, Sara Papenbrock, Anne Hildebrand, Elilarasi Ratnalingam, Eva Meisenzahl, Franziska Herrmann, Susanne Meinert, Elisabeth Leehr, Julia Hubbert, Judith Krieger, Hannah Meinert, Theresa Slump, Igor Nenadic, Andreas Jansen, Nooshin Javaheripour, Florian Thomas-Odenthal, Hamidreza Jamalabadi, Benjamin Straube, Marco Hermesdorf, Maike Richter, Raimund Helbok, Xiaoyi Jiang, Nils Opel, Klaus Berger, Tilo Kircher, Udo Dannlowski, Tim Hahn, Nils R. Winter, Ramona Leenings

**Affiliations:** Institute for Machine Learning in Medicine with focus area psychiatry; Institute for Translational Psychiatry, University of Münster, Germany; Institute for Geoinformatics, University of Münster, Germany; Department of Psychiatry and Neuroscience, Charité Universitätsmedizin Berlin, Berlin, Germany; German Center for Mental Health, Berlin, Germany; Institute of Epidemiology and Social Medicine, University of Münster, Germany; Department of Psychiatry and Psychotherapy, University of Marburg, Marburg, Germany; Center for Mind, Brain and Behavior, University of Marburg, Marburg, Germany; Department of Psychiatry, Medical School and University Medical Center OWL, Protestant Hospital of the Bethel Foundation, Bielefeld University, Bielefeld, Germany; German Center for Mental Health, Jena-Magdeburg-Halle, Germany; Center for Intervention and Research on Adaptive and Maladaptive Brain Circuits Underlying Mental Health, Jena-Magdeburg-Halle, Germany; Faculty of Mathematics and Computer Science, University of Münster, Münster, Germany; Department of Neurology, Kepler University Hospital, Johannes Kepler University Linz, Linz, Austria; Clinical Research Institute of Neurosciences, Kepler University Hospital, Johannes Kepler University, Linz, Austria; Medizinische Fakultät, Heinrich-Heine-Universität Düsseldorf, Düsseldorf, Germany

## Abstract

Major depressive disorder (MDD) has been associated with accelerated structural brain aging, yet whether this reflects a pre-existing neurobiological vulnerability, a dynamic acute state effect, or an accumulating biological residual remains unresolved. Across two longitudinal cohorts (N=3220), including a unique sample of 78 initially healthy individuals who transitioned into their first depressive episode during the study course, we systematically tested all three hypotheses. Patients with diagnosed MDD showed elevated MRI-derived brain age relative to healthy controls (1.4 and 2.5 years across cohorts). For the vulnerability hypothesis, individuals scanned prior to their first episode showed no baseline elevation, despite already demonstrating subclinical elevations in self-reported symptom severity, indicating that advanced brain age does not precede illness onset. For the state hypothesis, we found no acceleration of brain aging following the first depressive episode, and longitudinal brain age trajectories were independent of acute clinical symptom severity. Finally, neither episode duration nor recurrence scaled with brain age. Accelerated brain aging in depression is therefore neither an antecedent vulnerability nor an acute state marker of the first episode, but rather a stable biological feature of a long term illness course.

## Introduction

Major depressive disorder (MDD) represents a global health challenge of the modern era (1). Its course is often characterized by recurrence, chronicity, and accumulating functional impairment (2,3). How this cumulative history is encoded, and potentially reinforced by, brain structure remains an open question. Structural changes have been reported as cortical thinning in the frontal and temporal lobes and as gray matter volume reductions in subcortical structures such as the hippocampus (4–6), yet these effects are typically subtle and spatially distributed, and findings often vary across samples (7–11). In recent years, the brain age paradigm has therefore gained considerable traction as a promising complementary approach to quantify the impact of the disease (12).

Brain age models are trained to predict chronological age from structural MRI in healthy cohorts, thereby establishing a normative model of structural brain aging across the human lifespan (13,14). The resulting model can then be used to predict the brain age in unseen samples (14). The discrepancy between an individual’s brain age and their chronological age is termed the Brain Age Gap (BAG) and serves as a summary of brain-structural health, where positive values are interpreted as accelerated aging (15,16).

In MDD, large-scale consortia have consistently identified a significant BAG elevation (17,18). However, evidence on the underlying mechanisms is conflicting. One line of evidence frames accelerated brain aging as a pre-existing liability, showing that elevated BAG correlates with increased suicide propensity and poorer treatment outcomes (19). In contrast, other studies suggest that these deviations are dynamic and state-dependent. Research indicates that the BAG may be acutely increased near illness onset and correlates with symptom burden and episode related cognitive impairment (20,21). Furthermore, multi-omics evidence suggests that clinical remission may even diminish systemic aging acceleration (6). However, this state interpretation remains controversial, as multi-site consortium analyses report only modest coupling between BAG and current symptom severity, arguing against a purely state-like marker (17,18). As a third line of argumentation, the cumulative residual hypothesis posits that each period of depressive burden leaves behind a neurobiological residual, which progressively accumulates over time. Supporting this concept, elevated BAG in chronic and recurrent disease states predicts future depressive recurrences (22) and correlates with key metrics of disease progression, such as total duration of illness (17,23). This points to a lasting neurobiological residual that extends beyond the potential artifacts of long-term pharmacological exposure (23).

The primary obstacle to resolving these conflicting reports lies in the design of existing studies: cross-sectional data cannot disentangle whether this structural deviation precedes the illness or emerges from it. Resolving this temporal sequence requires longitudinal study designs that track the transition from health to disease. Capturing individuals scanned both before and after their first depressive episode (converters) is inherently challenging and such samples remain scarce in the literature (24).

Here, we leverage two independent, large-scale longitudinal datasets of N=3,220 individuals encompassing a rare subsample of n = 78 individuals who converted from Healthy Control (HC) to MDD during the study course. Within this dual-cohort framework, we systematically tested three competing temporal hypotheses:

- Hypothesis 1 (Vulnerability): Converters already exhibit an elevated BAG at baseline, prior to their first depressive episode, indicating a pre-existing structural deviation from expected age.
- Hypothesis 2 (State): The transition into a first episode triggers an immediate increase in BAG, and BAG fluctuates with acute clinician-rated and self-reported symptom severity.
- Hypothesis 3 (Residual): An elevated BAG in MDD is driven by the cumulative residual impact of illness duration and previous episodes, with each episode contributing an additive biological burden.

## Methods

### Participants and Study Design

Data for this study were drawn from independent, longitudinal neuroimaging cohorts: the Marburg-Münster Affective Disorders Cohort Study (MACS), the Münster Neuroimaging Cohort (MNC), and the BiDirect Study. Due to comparable demographic distributions and harmonized study protocols, participants from MACS and MNC were combined to form our Discovery Dataset. The independent BiDirect cohort served as a designated Replication Dataset, because its participants were on average 14 years older at baseline (see Supplementary Table S1) and followed different study protocols. All participants provided written informed consent, and the studies were approved by the local ethics committees. Participants in MACS and MNC received financial compensation for their time.

From an initial pool of 3,220 participants with multiple follow-up measurements (2 for MACS, up to 8 for MNC and up to 6 for BiDirect), we applied a multi-step quality control pipeline to ensure data integrity. Assessments were excluded if demographic records (age or sex) were incomplete, if the MRI scan was missing, or if brain age prediction failed (Supplementary Table S2). This resulted in a high-quality longitudinal sample of 6,122 scans from 2,718 participants with complete demographic data and successful brain age estimation (Supplementary Table S1).

From this quality-controlled pool, balanced cohorts were sub-selected for downstream longitudinal analyses using nearest-neighbour matching. Participants were initially recruited as either healthy controls (HC) or patients with diagnosed MDD. However, over the course of the ∼2-year longitudinal follow-up, a subset of the baseline healthy sample incidentally transitioned to a first depressive episode. These individuals (n = 78) were categorized as the converter group (CON), defined by the absence of any psychiatric diagnosis at baseline and fulfillment of DSM IV criteria for MDD at follow-up assessment.

Detailed information on the study samples, including inclusion criteria and methods, is provided in the corresponding publications (MACS: Kircher et al., Vogelbacher et al. (25,26); MNC: Dannlowski et al., Opel et al. (27,28); BiDirect: Teismann et al. and Wulms el al. (29,30))

### Clinical Assessments

#### Discovery Dataset

The presence or absence of psychiatric diagnoses at baseline and follow-up was verified by trained personnel using the Structured Clinical Interview for DSM-IV-TR (SCID-IV) (31). A retrospective assessment according to the life-chart method (32) was administered at the clinical follow-ups to capture the individual longitudinal disease courses, and was used to identify the transition to MDD in the converter group. Current depression severity was quantified using both the clinician-administered Hamilton Depression Rating Scale (HAMD) (33) and the self-reported Beck Depression Inventory (BDI) (34). At each clinical follow-up, episode recurrence status (single vs. recurrent) was determined based on whether an individual experienced >= 1 new depressive episode during the inter-assessment interval, and the cumulative lifetime duration of depressive episodes was recorded as a self-reported estimate in months. To account for the potential confounding effects of psychotropic medication in the Discovery dataset, a composite medication load index was calculated for each individual based on dose and variety of medications taken, integrating periods with and without medication during the follow-up interval (for detailed calculation, see Supplementary Methods).

#### Replication Dataset

Psychiatric diagnoses were evaluated using the German version of the Mini-International Neuropsychiatric Interview 5.0.0.0 (35), based on DSM-IV criteria. Conversion to MDD was defined by participants’ self-reported first physician diagnosis of depression since their previous study visit. Current depression severity was quantified at all timepoints using the self-reported Center for Epidemiologic Studies Depression (CES-D) Scale (36). Detailed psychotropic medication status was not recorded for the Replication dataset.

### MRI Acquisition

Structural T1-weighted high-resolution anatomical images were acquired in all cohorts on 3-Tesla MRI scanners using three-dimensional fast gradient echo sequences. Detailed sequence parameters for each scanner are provided in the Supplementary Methods. In the MACS cohort, data were acquired at two sites (Marburg and Münster). In the MNC and BiDirect cohort, data were collected on a single 3T scanner in Münster. To mitigate the sensitivity of brain age models to hardware variations, all scanner site configurations (MACS Marburg, MACS Münster, MNC Münster and BiDirect Münster) were explicitly controlled for in all downstream statistical models.

### Brain Age Prediction

Brain age predictions were extracted using the brainageR software developed by Cole et al. (37–39), which utilizes a Gaussian Processes regression to predict brain age directly from raw, T1-weighted NIfTI formatted MRI scans. For our distributed prediction pipeline, we utilized a containerized version of the software accessible on https://github.com/fprados/brainageR_dockerfile. Total Intracranial Volume (TIV) was also extracted from brainageR’s SPM segmentation for use as a covariate in downstream analyses.

The BAG was calculated as the difference between the predicted brain age and the participant’s chronological age.

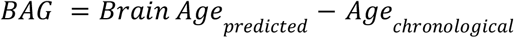

To account for the age-related bias inherent in raw BAG estimates (40,41), we included age as a covariate, in line with recommended practice (41–43).

### Data Matching and Covariate Control

Given the small expected effect sizes in psychiatric neuroimaging (44,45) and the sensitivity of brain age models to hardware variations across multi-site data (46,47), we implemented a rigorous matching protocol to isolate the neurobiological effects of depression onset (48). We matched each CON with 5 HC or MDD subjects to increase statistical power while maintaining acceptable covariate balance. To standardize longitudinal evaluation across the sample, we restricted our analyses to a maximum of two timepoints per subject by selecting baseline and primary follow-up assessments for the control and MDD groups, and the two scans directly flanking first episode onset for converters.

Following the matching protocol, the final longitudinal samples were composed of 858 individuals with 1487 scans (see Table 1). These matched sub-samples ensure that subsequent comparisons reflect clinical trajectories rather than artifacts of scanner site or baseline demographic imbalance. Comprehensive statistical validation of the matching procedure is detailed in the Supplementary Results.

**Table 1.** Demographic and clinical characteristics across the two datasets after age sex matching at Baseline.

|  | HC |  | MDD |  | CON |  |
| --- | --- | --- | --- | --- | --- | --- |
| **Discovery (n=517)** | n | mean (±SD) | n | mean (±SD) | n | mean (±SD) |
| Total | 235 |  | 235 |  | 47 |  |
| Age |  | 35.42 (12.29) |  | 34.62 (12.63) |  | 34.73 (11.70) |
| Female Sex | 127 |  | 126 |  | 25 |  |
| Scanner Site* | 98/31/106 |  | 103/32/100 |  | 21/6/20 |  |
| Follow-Up Scans | 168 |  | 182 |  | 32 |  |
| HAM-D |  | 0.94 (1.48) |  | 12.00 (7.89) |  | 1.94 (2.51) |
| BDI |  | 3.07 (3.41) |  | 21.81 (11.44) |  | 6.23 (6.03) |
| **Replication (n=341)** | n | mean (±SD) | n | mean (±SD) | n | mean (±SD) |
| Total | 155 |  | 155 |  | 31 |  |
| Age |  | 52.42 (8.43) |  | 52.18 (8.11) |  | 52.40 (8.57) |
| Female Sex | 95 |  | 92 |  | 19 |  |
| Follow-Up Scans | 125 |  | 97 |  | 25 |  |
| CES-D |  | 7.76 (5.59) |  | 25.25 (13.21) |  | 13.32 (6.36) |
| *three scanner sites are: MACS Marburg, MACS Münster and MNC Münster |  |  |  |  |  |  |

### Statistical Analyses

All statistical analyses were conducted in R (v.4.5.1). Linear mixed-effects (LME) models were implemented using the *lmerTest* package, with ANCOVA computed via the *car* package. For all inferential models, the raw, uncorrected BAG served as the dependent variable. To rigorously correct for established age-prediction biases within the model, chronological age (both linear and quadratic terms) was included as a covariate in all analyses. To account for demographic and clinical heterogeneity, all hypotheses were tested independently within the Discovery and Replication datasets. Post-hoc pairwise contrasts and marginal trends were calculated using estimated marginal means via the *emmeans* package.

#### Baseline Comparison (Vulnerability Hypothesis)

To test for pre-existing neurobiological and clinical deviations prior to illness onset, we conducted cross-sectional analyses of covariance (ANCOVAs) using baseline data, with group status (HC, CON, MDD) as the independent variable. Separate models were run for BAG (to test the vulnerability hypothesis) and clinical severity measures (BDI, HAM-D, or CES-D; for sample characterization). Pairwise group differences were evaluated using two-tailed Tukey’s post-hoc corrections. Analyses prioritized the primary, hypothesis-driven contrast evaluating whether advanced brain aging precedes depression onset (CON > HC), while the secondary contrast (MDD > HC) served to validate established literature effects within our sample. All models were adjusted for baseline chronological age (linear and quadratic), biological sex, TIV, and, where applicable, scanner site. To assess the robustness of our findings, sensitivity analyses were conducted by re-estimating the ANCOVAs with BMI (Body Mass Index) and medication load included as additional covariates to confirm that primary group contrasts remained stable. To benchmark the group-level differences observed in our neuroanatomical approach, parallel ANCOVAs were applied to baseline clinical severity scores (BDI, HAM-D, and CES-D) as outcome variables, adjusting for biological sex and chronological age (linear and quadratic).

#### Clinical State Dynamics (State Marker Hypothesis)

To evaluate whether advanced brain aging functions as a dynamic marker of symptomatic vs non-symptomatic states, we analyzed the association between BAG and depressive states using both longitudinal and cross-sectional modeling. First, the *Group* × *Time* interaction was assessed to capture the transition into a first depressive episode, comparing the temporal trajectories of converters and healthy controls. Fixed effects included group, time since baseline, chronological age terms, sex, and TIV. Random intercepts were specified for subjects and scanner sites. Pairwise contrasts of the groups’ estimated marginal temporal trends (slopes) were calculated post-hoc. Second, we isolated the coupling of BAG with acute symptom severity in the clinical subgroups (CON and MDD). An ANCOVA associated current clinical severity (BDI, HAM-D, or CES-D) to BAG at follow-up, adjusting for the chronological age (both linear and quadratic terms), sex, TIV and scanner site. In an additional change score model we captured longitudinal shifts by regressing the change in BAG between baseline and follow-up (ΔBAG) against the change in symptom severity (ΔBDI, ΔHAM-D or ΔCES-D), adjusting for baseline chronological age terms, sex, TIV, and scanner site.

#### Cumulative Disease Burden (Residual Hypothesis)

To test whether the magnitude of the aging effect is associated with cumulative disease burden, we applied an LME model restricted to the clinical populations (CON and MDD) in the discovery dataset. Disease burden was operationalized by incorporating either episode recurrence (categorical: healthy, no relapse, or relapse) or cumulative illness duration (defined as the total lifespan spent in depressive episodes in years) as fixed effects. The model is adjusted for age terms, sex, and TIV, with random intercepts for subject and scanner site. For the categorical recurrence analysis, post-hoc pairwise contrasts of the estimated marginal means were calculated to compare subgroups. As a sensitivity analysis to test for potential time-varying effects, we further extended both analyses with interaction terms between time and cumulative illness duration, as well as time and episode recurrence.

#### Equivalence Testing

To formally confirm the absence of clinically meaningful effects, Two One-Sided Tests (TOST) procedures were implemented across all models using the *TOSTER* package. Equivalence boundaries were configured according to the scale and clinical context of each specific hypothesis. For the baseline comparison prior to conversion, unstandardized equivalence boundaries were anchored to the baseline effect of established MDD in the respective cohort (±1.39 years in Discovery; ±2.51 years in Replication). For clinical symptom inventories we derived equivalence boundaries from established literature or distribution-based thresholds. These comprised a minimum clinically relevant change of ±7.0 points for clinician-rated HAM-D (49), a sample-specific threshold of ±3.87 points for self-reported BDI (17.5% of the MDD sample mean; (50)), and a half-standard-deviation margin of ±4.93 points for self-reported CES-D (51). For longitudinal aging slopes, change-score models, and continuous clinical correlations where explicit unstandardized cut-offs cannot be derived, standardized boundaries were adopted following consensus criteria for small effects (*d* = ±0.20 or *r* = ±0.10).

## Results

### The Vulnerability Hypothesis: Evaluating brain age pre-conversion

To evaluate whether accelerated brain aging represents a pre-existing state accumulated over the prior life course and indicating a vulnerability for depression, we analyzed cross-sectional BAG differences at the baseline assessment. At this timepoint, individuals in the CON group had no current psychiatric diagnosis and had not yet experienced their first depressive episode.

#### Cross-Sectional Baseline Brain Age Gap

The ANCOVA revealed a significant main effect of group on baseline BAG (*F(2, 507)* = 5.26, *p* = .006, 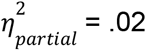; Supplementary Table S3), which was driven exclusively by the established illness: the MDD group showed a significantly higher BAG compared to HC at baseline (estimate = 1.39 years, *SE* = 0.44, *t(507)* = -3.14, *d* = 0.29, *p*_*Tukey*_ = .005). Sensitivity analyses revealed that the observed group difference in BAG for HC vs. MDD remained significant after controlling for BMI and medication load (p = .008). Neither BMI (*p* = .955) nor medication index (*p* = .191) were significant predictors of BAG in the full model. With regard to the vulnerability hypothesis, we found no evidence of BAG elevation in the CON group prior to disease onset. In contrast to the 1.39 year difference between HC and MDD, the brain age of HC and converters was nearly identical (estimate = 0.03 years, *SE* = 0.76, *t(507)* = -0.04, *d* = 0.006, *p*_*Tukey*_ > .99; see Figure 1A). To test whether this null finding reflects a true absence of a pre-onset structural shift, we conducted an equivalence test. Using the baseline MDD effect size (±1.39 years) as the equivalence margin, the HC vs. converter contrast was significantly equivalent (*t(507)* = 1.79, *p*_*TOST*_ = .037; 90% *CI* [-1.28, 1.22] years).

**Figure 1.**
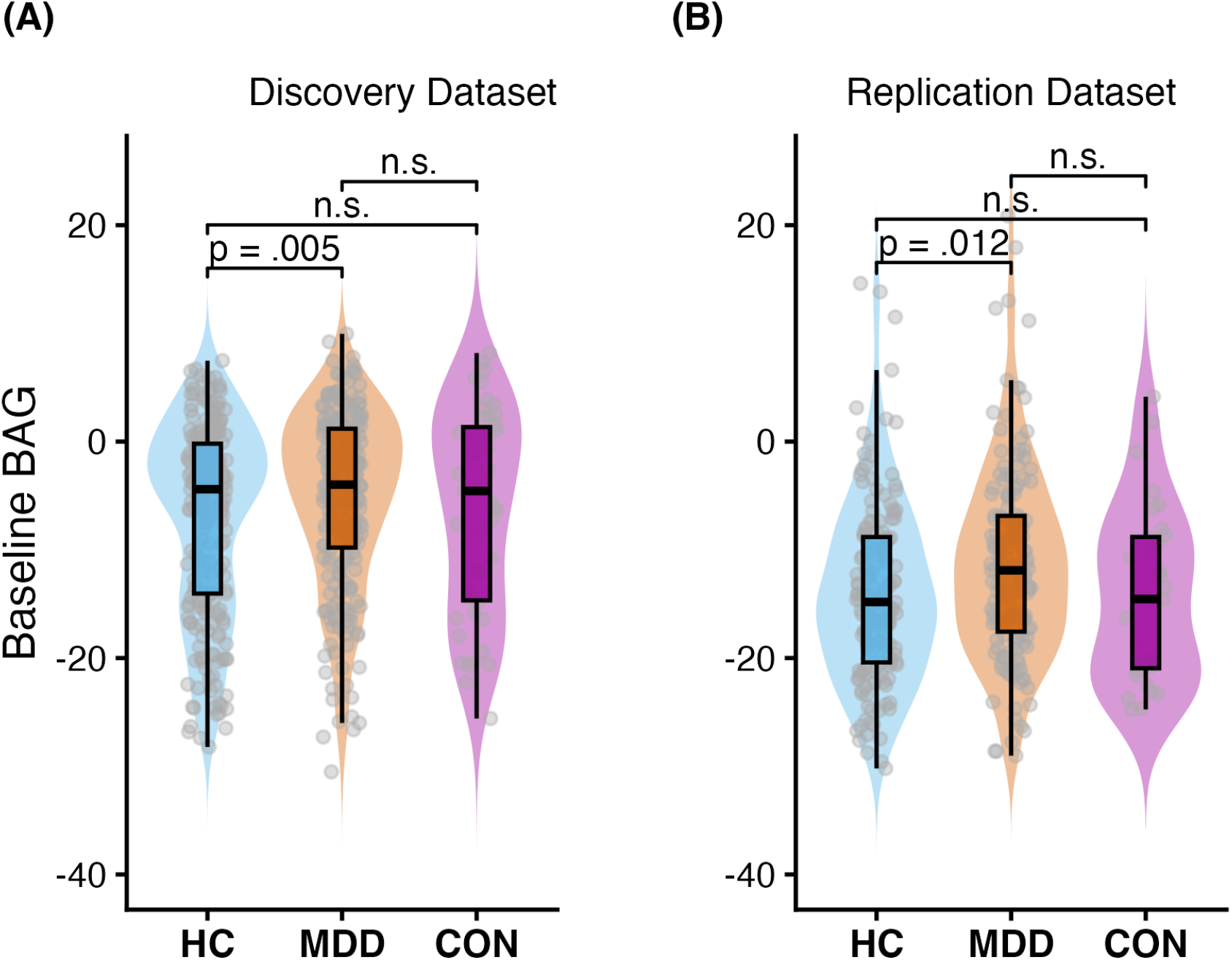
Comparison of BAG (brain age gap) between HC (healthy control) and CON (converter before first depressive episode) at baseline reveals no significant difference. HC and MDD (major depressive disorder) have a significantly different BAG at baseline in both datasets.

In the Replication dataset, these results were confirmed: we again observed a significant main effect of group on baseline BAG (*F(2, 334)* = 4.49, *p* = .012, 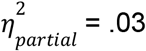). Similarly, the only significant post-hoc contrast was between HC and MDD (estimate = 2.51 years, *SE* = 0.88, *t(334)* = -2.86, *d* = 0.326, *p*_*Tukey*_ = .012; see Figure 1B, Supplementary Table S4). The baseline brain age gap between HC and CON was virtually identical (estimate = 0.03 years, *SE* = 1.52, *t(334)* = 0.02, *d* = 0.001, *p*_*Tukey*_ > .99). An equivalence test using the replication baseline disease effect (±2.51 years) as boundary approached formal statistical significance (*t(334)* = -1.64; *p*_*TOST*_ = .051; 90% *CI* [-2.47, 2.53] years).

#### Pre-Conversion Subclinical Symptom Profiles

To complement these neuroanatomical findings, we evaluated whether the converter group already showed elevated depressive symptoms before conversion, despite not yet meeting diagnostic threshold. The clinician-rated HAM-D showed a robust main effect of group (*F(2, 511)* = 253.76, *p* < .001, 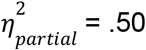). This effect was mainly driven by the MDD group, as post-hoc pairwise comparisons failed to differentiate the CON from the HC group (estimate = -1.01, *SE* = 0.88, *t(511)* = -1.16, *d* = -0.19, *p*_*Tukey*_ = .478). An equivalence test confirmed that the baseline clinical profiles of the HC and CON were statistically bounded within the pre-defined ±7-point threshold (*t(511)* = 12.56; *p*_*TOST*_ < .001; 90% *CI* [-0.43, 2.45] HAM-D Points).

Conversely, self-reported inventories captured sub-clinical phenotypes in the pre-clinical phase. Baseline differences between HC and CON subjects demonstrated a marginal trend on the BDI (*F(2, 496)* = 309.22, *p* < .001, 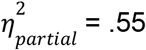; post-hoc: estimate = -3.17, *SE* = 1.36, *t(496)* = -2.32, *d* = -0.39, *p*_*Tukey*_ = .054). An equivalence test using the pre-defined threshold of ±3.87 points indicated that the two groups could not be considered equivalent (*t(496)* = -0.52; *p*_*TOST*_ = .303; 90% *CI* [0.93, 5.41] BDI points). The comparison was statistically significant for the CES-D in the Replication cohort (*F(2, 334)* = 122.72, *p* < .001, 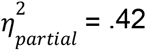; post-hoc: estimate = -5.56, *SE* = 1.94, *t(334)* = -2.87, *d* = -0.56, *p*_*Tukey*_ = .012). Applying the pre-defined ±4.93-point equivalence margin, the equivalence test was clearly non-significant (*t(334)* = 0.33; *p*_*TOST*_ = .628; 90% *CI* = [2.36, 8.76] CES-D points).

These findings suggest that BAG is not advanced prior to the onset of the first depressive episode, even though subclinical depressive symptoms likely exist. At the same time the increase in brain age in the MDD group (∼1.4 to 2.5 years) demonstrates that an established diagnosis is associated with a detectable deviation in brain age.

### The State Marker Hypothesis: Evaluating longitudinal brain age

To test whether biological brain aging functions as a dynamic state marker of depression, we examined whether BAG fluctuates with acute disease activity over time.

#### Longitudinal Brain Aging Trajectories

We tested whether the transition into a first depressive episode coincided with an acute acceleration of biological brain aging by employing a LME model, evaluating the interaction between group (HC, MDD, CON) and time (measured as years from baseline). This revealed no significant group-by-time interaction (*F(2, 388.19)* = 0.53, 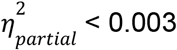, *p* = .588; see Supplementary Table S5), indicating that the change in BAG in-between two timepoints did not differ significantly between individuals transitioning to a first episode of MDD, patients with MDD, and sustained HC. Pairwise comparisons of the aging trajectories confirmed this longitudinal stability. The rate of brain aging in-between the baseline and follow-up timepoint in the CON groups was statistically indistinguishable from the trajectories of HC (estimate = 0.05 years/year, *SE* = 0.12, *t(387)* = 0.45, *d* = 0.04, *p*_*Tukey*_ = .894) and the MDD group (estimate = -0.01 years/year, *SE* = 0.12, *t(386)* = -0.08, *d* = -0.01, *p*_*Tukey*_ = .996; see Figure 2A, Supplementary Table S6). To formally confirm the absence of acute trajectory divergence, equivalence tests were conducted against a standardized small-effect boundary (*d* = ±0.20, corresponding to a raw margin of ±0.250 BAG years/year). The longitudinal rate of brain aging in the CON group was statistically equivalent to that of HC (*t(387)* = -1.66, *p*_*TOST*_ = .048; *90% CI* = [-0.14, 0.25] years/year) as well as established MDD patients (*t(386)* = 1.98, *p*_*TOST*_ = .024; *90% CI* = [-0.21, 0.19] years/year).

**Figure 2.**
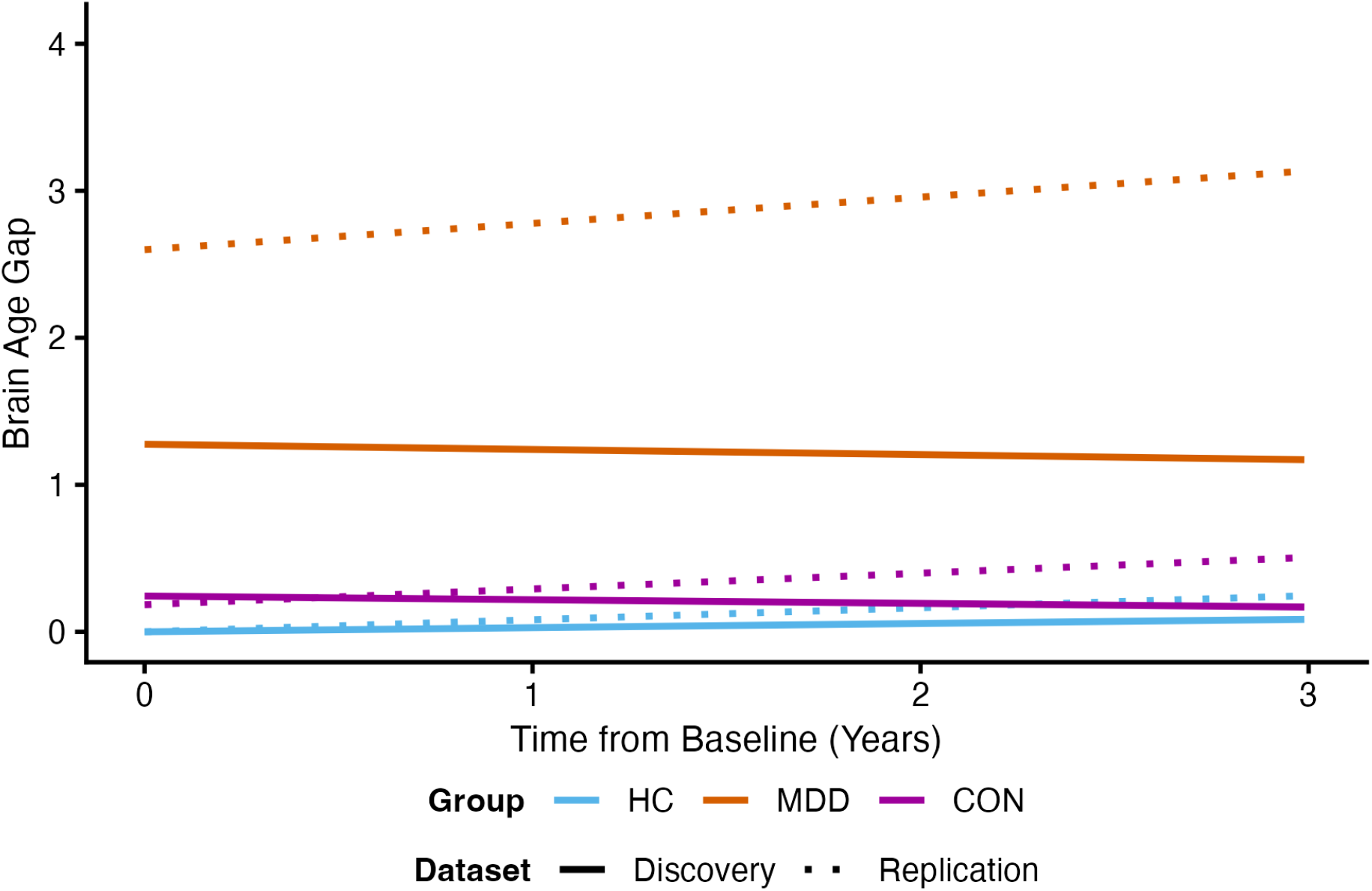
Longitudinal trajectories of the covariate-adjusted Brain Age Gap (BAG) across independent cohorts. Modeled trajectories are displayed by group (HC = Healthy Control, CON = Converter, MDD = Major Depressive Disorder) for the independent datasets. Displayed lines represent predicted values derived from longitudinal linear mixed-effects models adjusting for chronological age (linear and quadratic terms), biological sex, total intracranial volume, and MRI scanner. To facilitate direct visual comparison of relative deviations across cohorts, BAG estimates were baseline-centered by anchoring the HC group at zero. No significant group-by-time interactions were observed in either dataset, despite a 17 years higher average age in the replication data.

In the Replication dataset these results were fully replicated. We observed no significant group-by-time interaction (*F(2, 249.21)* = 0.33, 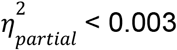, *p* = .720) and no significant pairwise differences in aging trajectories (see Figure 2B, Supplementary Table S6). The annual rate of brain aging in the CON group remained statistically indistinguishable from both HC (*estimate* = -0.025 years/year, *SE* = 0.190, *t(249)* = -0.13, *p*_*Tukey*_ = .990) and established MDD patients (*estimate* = 0.072 years/year, *SE* = 0.191, *t(249)* = 0.37, *p*_*Tukey*_ = .926). Mirroring the discovery protocol, equivalence tests using the standardized small-effect boundary (*d* = ±0.20, corresponding to a raw margin of ±0.422 BAG years/year) formally confirmed trajectory equivalence for both the HC comparison (*t(249)* = 2.09, *p*_*TOST*_ = .019; *90% CI* = [-0.339, 0.289] years/year) and the MDD comparison (*t(249)* = -1.83, *p*_*TOST*_ = .034; *90% CI* = [-0.244, 0.387] years/year).

#### Symptom Severity and Brain Age Gap Correlation

As an exploratory additional analysis within the established MDD subgroup, we evaluated whether concurrent depression severity scales with absolute brain aging at the follow-up assessment. No significant association was observed between BAG magnitude and clinician-rated symptom severity (HAM-D: *F(1)* = 0.004, p = .950, 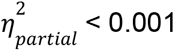) or self-reported depression severity (BDI: *F(1)* = 0.095, p = .759, 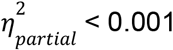). The partial correlations were negligible for both metrics (HAM-D: *r* = -0.004, BDI: *r* = 0.022). Equivalence tests on the partial correlations against a standardized consensus boundary of r = ±0.10 yielded statistically indeterminate results for both HAM-D (z = 1.37, *p*_*TOST*_ = .085, *90% CI =* [-0.12, 0.11]) and BDI (z = -1.07, *p*_*TOST*_ = .141, *90% CI* = [-0.10, 0.14]). Similarly, analyses within the Replication dataset yielded no significant effects for self-reported clinical scores (CES-D: *F(1)* = 0.888, *p* = .348, 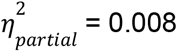, *r* = -0.087). The equivalence test against the same boundary (r = ±0.10) was likewise statistically indeterminate (z = 0.14, *p*_*TOST*_ = .444, *90% CI* = [-0.24, 0.07]). Together, these results suggest that the observed BAG is independent of current symptomatic state (see Figure 3, Supplementary Table S7).

**Figure 3.**
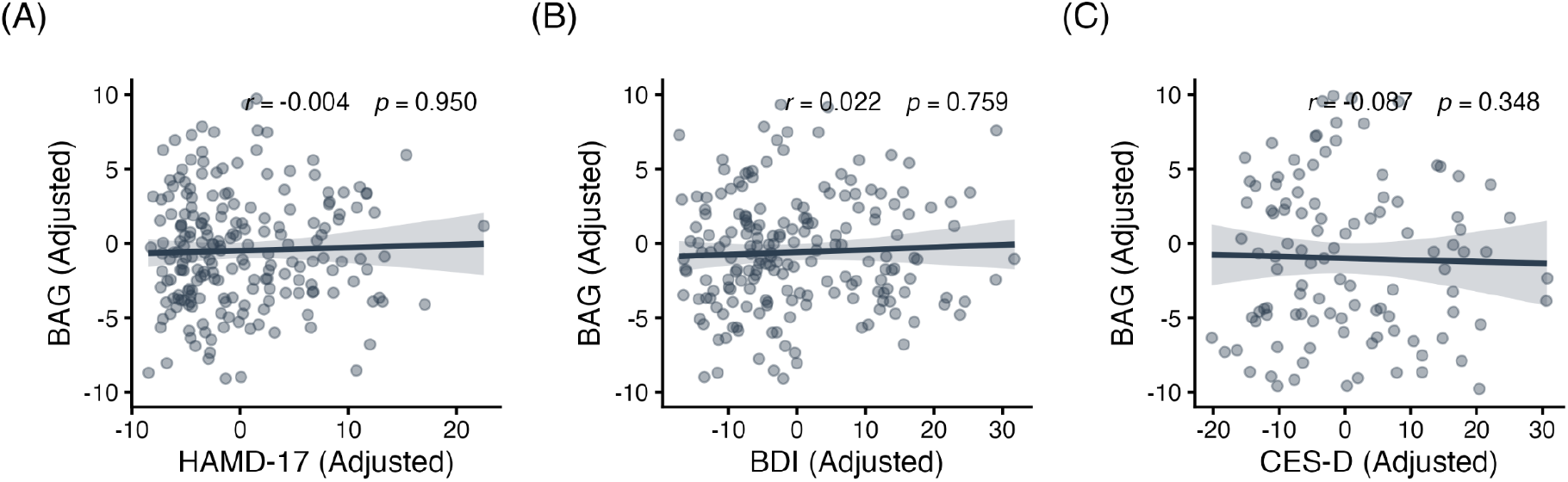
Association between the Brain Age Gap (BAG) and symptom severity at follow-up. (A) clinician-rated HAM-D and (B) self-reported BDI in the Discovery dataset (C) self-reported CES-D in the Replication dataset. All data shown is adjusted for age, age^2^, sex, total intracranial volume, and scanner site to visualize the isolated relationship between disease severity and BAG. Regression lines are shown with 95% confidence intervals.

#### Relationship Between Longitudinal Symptom Changes and Brain Aging

As the dichotomous change in disease-status was not associated with increased brain aging at follow-up, we instead asked whether continuous changes in symptom severity between timepoints were coupled to changes in BAG. We found that longitudinal changes in brain aging were independent of the clinical course of the disease. Change score models revealed no significant association between ΔBAG and changes in either clinician-rated symptom severity from baseline to follow-up (ΔHAM-D: *F(1)* = 0.001, p = .973, 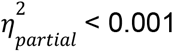), or self-reported depression scores (ΔBDI: *F(1)* = 0.051, p = .822, 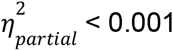) and negligible partial correlations (ΔHAM-D: *r* = -0.002, ΔBDI: *r* = 0.017). Equivalence tests against a standardized small-effect boundary (r = ±0.10) were statistically indeterminate for both metrics (ΔHAM-D: z = -1.39, *p*_*TOST*_ = .082, *90% CI* = [-0.12, 0.11]; ΔBDI: z = -1.13, *p*_*TOST*_ = .129, *90% CI* = [-0.10, 0.14]). In the Replication dataset the self-reported CES-D scores showed only a marginal trend (ΔCES-D: *F(1)* = 3.065, p = .083, 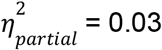, r = 0.161), with the point estimate falling outside the equivalence margin (z = 0.66, *p*_*TOST*_ = .746, *90% CI* = [0.01, 0.31]). Taken together, these findings indicate no robust relationship between longitudinal shifts in depressive symptomatology and MRI-derived brain age metrics (see Figure 4, Supplementary Table S8).

**Figure 4.**
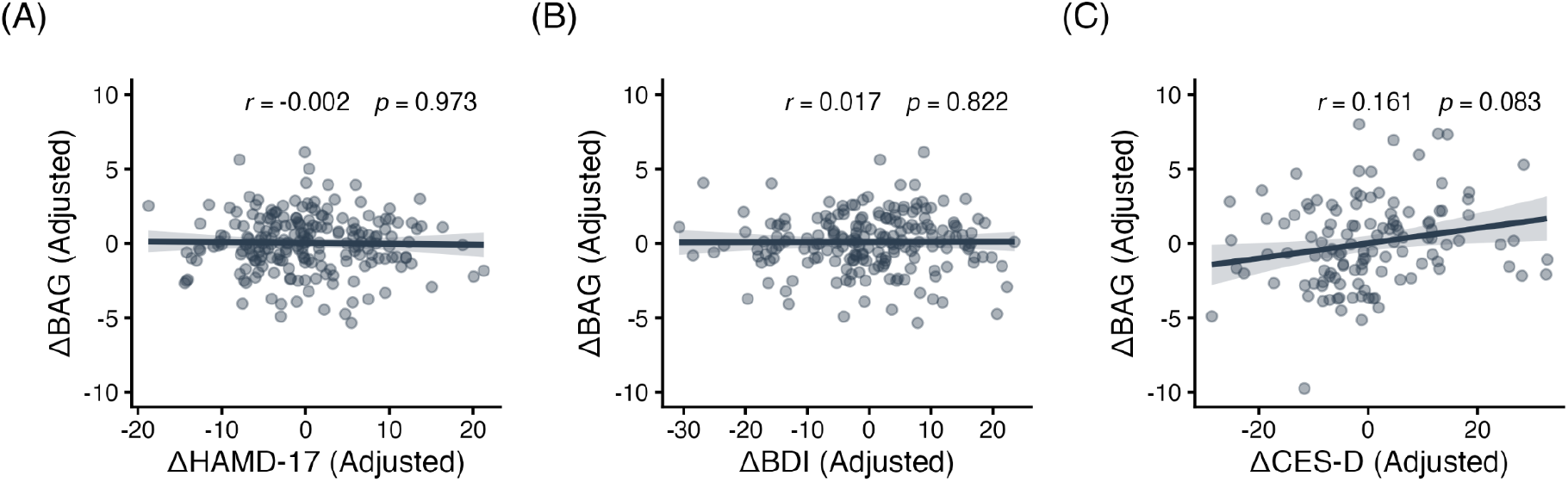
Association between change in brain aging (ΔBAG) and change in symptom severity from baseline to follow-up. (A) ΔHAM-D and (B) ΔBDI in the Discovery dataset, (C) ΔCES-D in the Replication dataset. All data shown is adjusted for age, age^2^, sex, total intracranial volume, and scanner site to visualize the isolated relationship between disease severity and BAG. Regression lines are shown with 95% confidence intervals.

### The Residual Hypothesis: Evaluating brain age as a marker of cumulative disease burden

In our final analysis, we tested whether accelerated brain aging functions as a cumulative consequence of disease burden in MDD patients. Within the symptomatic groups (MDD and CON), we examined the relationship between BAG and two markers of lifetime illness progression: cumulative episode duration and number of episodes.

#### Cumulative Illness Duration and Structural Aging Progression

We found that the cumulative lifetime duration of depressive episodes was not a significant predictor of brain aging (*estimate* = -0.005, *t*(270.8) = -1.535, *p* = .126; see Supplementary Table S9). The partial effect size was negligible 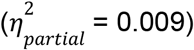, particularly relative to chronological age (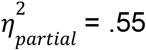, *p* < .001). The partial correlation was likewise negligible (r = -0.093). An equivalence test against the standardized boundary of r = ±0.10 was statistically indeterminate (*z* = 0.12, *p*_*TOST*_ = .453, *90% CI* = [-0.19, 0.01]). A sensitivity analysis revealed no significant interaction with time (estimate = -0.001, *t(225.2)* = -0.705, *p* = .482), demonstrating that historical disease burden does not accelerate the trajectory of brain aging. The corresponding partial correlation was trivial (*r* = -0.047), and the equivalence evaluation was similarly indeterminate (*z* = 0.80, *p*_*TOST*_ = .213, *90% CI* = [-0.16, 0.06]). Notably, the variance components analysis revealed that between-person variance (25.55) substantially exceeded within-person longitudinal variance (1.85). This indicates a high degree of within-person stability in aging progression across the longitudinal observation window.

#### Brain Aging Across Single vs. Recurrent Episode Courses

Comparing individuals with single versus multiple depressive episodes, we found that the frequency of depressive episodes did not significantly modulate biological brain aging (*F*(2, 273.3) = 0.032, *p* = .969, 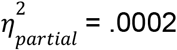; see Figure 5, Supplementary Table S10). Post-hoc pairwise comparisons revealed that BAG was statistically indistinguishable between relapsing and non-relapsing patients (*estimate* = -0.009, *t(302.9)* = -0.029, *p*_*Tukey*_ = .999). An equivalence test using a standardized boundary of d = ±0.20 confirmed that this difference is statistically equivalent to zero (*t(302.9)* = -3.52, *p*_*TOST*_ < .001, *90% CI* = [-0.54, 0.52]). BAG was similarly indistinguishable between non-relapsing cases and the pre-clinical state of converters (*estimate* = 0.085, *p* = .968; see Supplementary Table S11), which also show no difference in equivalence tests using a boundary of d = ±0.20 (*t(244.5)* = -2.89, *p*_*TOST*_ = .002, *90% CI* = [-0.49, 0.66]). Again, the longitudinal trajectories revealed no significant interaction with Time (estimate = -0.148, *t(245.5)* = 0.955, *p* = .340), indicating that the rate of brain aging over time did not diverge between single and multiple-episode courses. The partial correlation for this interaction was trivial (*r* = 0.061), and an equivalence test against a standardized limit of r = ±0.10 was statistically indeterminate (*z* = -0.62, *p*_*TOST*_ = .269, *90% CI* = [-0.044, 0.165]).

**Figure 5.**
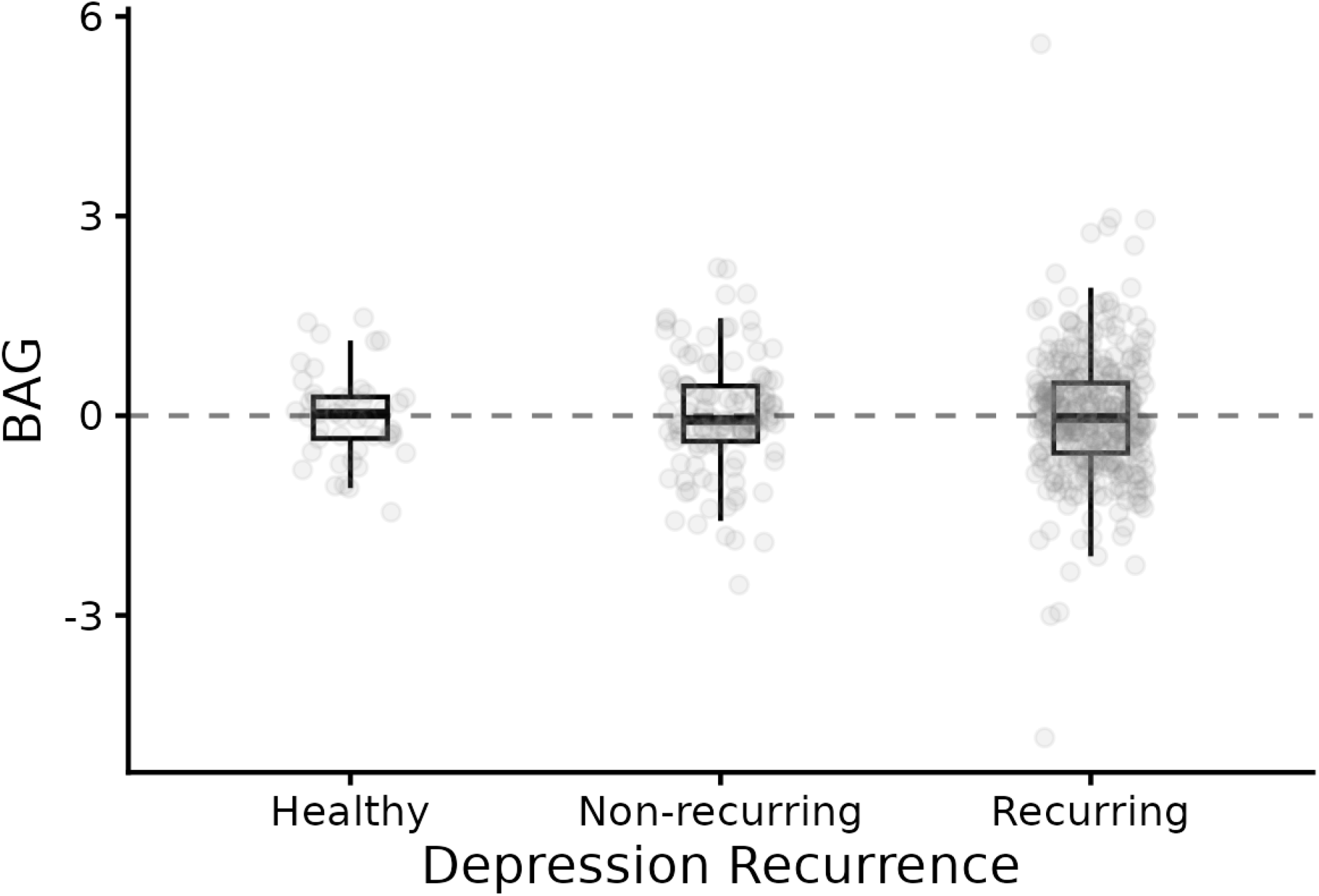
Biological brain aging across clinical progression stages of major depressive disorder. The distribution of the Brain Age Gap (BAG) across distinct milestones of disease course. To isolate the unique effect of disease recurrence, the vertical axis represents residuals from a linear mixed-effects model adjusted for chronological age, age^2^, biological sex, total intracranial volume, scanner site, and subject-level random intercepts. Linear mixed-effects regression revealed no significant structural divergence across these clinical stages (p = .976).

These results show no association of clinical progression and MRI derived brain age, arguing against the cumulative residual model in this cohort.

## Discussion

The primary objective of this study was to elucidate whether accelerated brain aging in patients with Major Depressive Disorder represents a pre-existing neurobiological susceptibility, an acute state marker, or an accumulated biological residual of illness burden. By comparing a cohort of healthy individuals who subsequently transitioned to MDD with matched sustained HC and established MDD patients, we found no evidence of accelerated brain aging prior to disease onset. The observed difference between the HC and MDD groups is in line with current research (17,18,22). Furthermore, longitudinal brain aging trajectories did not accelerate following illness onset and remained uncoupled from both acute symptom severity and longitudinal symptom fluctuations. Finally, clinical indicators of lifetime illness burden (cumulative disease duration and episode recurrence) did not predict the magnitude of BAG.

Because our conclusions rest on limited sample sizes and non-significant results, we employed formal equivalence testing. We demonstrate that baseline pre-conversion differences, longitudinal aging trajectories across independent cohorts, and episode recurrence frequencies are all statistically bounded within a small-effect threshold (d < ±0.20). This formal equivalence moves our conclusions beyond a mere failure to reject the null hypothesis and supports the conclusion that biological brain aging remains largely stable across early disease stages.

Rather than an accumulating biological residual that scales linearly with ongoing illness, our findings point toward a state-independent structural phenotype that characterizes established MDD. It appears to behave like a step function: accelerated brain aging distinguishes established MDD from health but does not scale with ongoing illness burden thereafter.

A sensitivity analysis accounting for BMI and medication load argued against metabolic or pharmacological explanations. Notably, MDD diagnosis remained a significant predictor of BAG even after these factors were included in the model and neither BMI nor medication load significantly predicted biological aging in our sample.

Across our samples, BAG was not significantly correlated with BDI, HAM-D, or CES-D scores. This supports conceptualising BAG as a marker of established disease rather than of acute symptom severity. This decoupling is consistent with recent evidence from Förster et al. (22), who similarly found that episode recurrence was not a predictor of BAG in MDD.

A parallel dissociation is well documented in multiple sclerosis (MS), where relapse activity and the underlying neurodegenerative process are partly independent and brain atrophy progresses during clinical remission by mechanisms distinct from the acute inflammatory episodes (52). If, as in MS, the structural process that elevates BAG runs separately from the symptomatic episodes captured by clinical scales, a brain age marker should distinguish patients from controls while remaining flat with respect to current severity and episode count. This is the pattern we observe.

However, the divergence among our baseline metrics reveals a clear decoupling of subjective distress, clinical signs, and structural neuroanatomy in the pre-conversion phase. While self-report inventories (BDI, CES-D) successfully captured an internal, sub-clinical prodromal state in future converters, both clinician-rated severity (HAM-D) and global brain structure (BAG) remained strictly equivalent to HC. This indicates that while self-reports are sensitive to early, internalizing cognitive distress, observable neurodegenerative signs and macrostructural brain changes have not yet materialized. This decoupling suggests that early subjective affective burden is insufficient to alter global structural brain trajectories, providing evidence against the vulnerability hypothesis. Instead, it reinforces the conceptualization of BAG as a stable, state-independent threshold phenotype that is absent during the prodromal phase and emerges only once the clinical disorder has consolidated.

One limitation of our studies lies in how MDD conversion was defined in the two cohorts underlying this study. In the Discovery dataset, converters were identified through structured clinical interviews and life-chart reconstructions, whereas the Replication dataset relied on self-reported physician diagnosis, an approach more susceptible to measurement noise. The absence of pre-conversion BAG elevation across both cohorts significantly strengthens the finding, showing that it persists under stringent and permissive criteria alike. Because of this more permissive definition, the Replication analyses carry less evidential weight and should be interpreted as consistent with, rather than independent confirmation of, the finding in the Discovery set.

In the evaluation of the cumulative residual hypothesis, measurement noise and metric granularity must also be considered. Cumulative episode duration relied on retrospective self-report, which is susceptible to recall bias and tends to attenuate observed association sizes. Furthermore, operationalizing disease burden via single clinical variables, such as duration or categorical recurrence, may under-represent the multidimensional complexity of clinical trajectories compared to data-driven composite scores (53,54). Thus the observed null relationship between lifetime burden and BAG could in part reflect imprecise or isolated burden metrics rather than a true absence of effect. However, the variance decomposition, which does not depend on retrospective recall and clinical operationalization, showed that between-person variance in BAG exceeded within-person variance more than tenfold (25.55 vs 1.85). This indicates that an individual’s brain age was largely fixed across the follow-up window, which does not support a model of accumulating biological residuals.

A primary methodological constraint of the present study is the sample size of the CON group (n = 78). The transition from a healthy baseline to a first major depressive episode is a rare event that cannot be experimentally induced. Such data can only be obtained through the prospective monitoring of large community-based cohorts, from which only a small proportion of individuals will incidentally convert. Furthermore, maintaining longitudinal retention in this population is challenging, as the onset of MDD often entails significant clinical and psychosocial upheaval. Consequently, while our CON group is modest in absolute terms, it provides a rare observational window into the neurobiological onset of depression.

In the present study, we interpreted the BAG as a global, age-agnostic marker consistent with most prior work. Emerging evidence (55), however, suggests that age-specific evaluations of the BAG can reveal age-specific meaning. Larger, well-powered samples may therefore reveal nuanced age- and sex-dependent effects that our cohort was not able to detect.

Moreover, it is important to consider that biological brain aging is a high-dimensional process that may not be fully captured by a single global index. Recent evidence suggests that brain aging unfolds along multiple, partially dissociable structural dimensions rather than a uniform trajectory (56). In psychiatric cohorts, subtle subtype-specific patterns are frequently obscured by conventional case-control contrasts and global metrics (57). Indeed, localized morphometric analyses provide evidence for such specific structural deviations, as Kraus et al. (58) used VBM to identify regionally increased gray matter volume in converters relative to both HC and MDD. Therefore, while our results demonstrate that the BAG remains remarkably constant across the early disease course, this stability does not preclude the existence of more nuanced, multivariate structural shifts. Future research should move beyond global measures to explore whether specific regional patterns accompany the transition into established MDD.

Overall, our results suggest that the post-onset window is a critical target for intensive longitudinal research. Because the elevated BAG phenotype is already present and stable in patients with established MDD, but does not emerge during the first active episode itself, future work must focus on the transition phase following first-episode recovery. Elucidating exactly when and how this structural gap emerges after the first major episode will be essential for refining the predictive and diagnostic utility of brain age markers in clinical psychiatry.

## Data Availability

The raw imaging and clinical datasets analyzed during the current study are not publicly available due to data privacy regulations and institutional governance restrictions. However, the data can be requested directly from the primary investigators and steering committees of the respective cohorts (e.g., MACS, MNC, BiDirect) upon formal data access application and approval.

## Funding

This work was funded by the IMF research instrument of the medical faculty of Münster (grant LE 1 1 24 09 to R.Leenings), the German Research Foundation (DFG, grant FOR2107 DA1151/5-1, DA1151/5-2, DA1151/9-1, DA1151/10-1, DA1151/11-1 to UD; SFB/TRR 393, project grant no 521379614) and the Interdisciplinary Center for Clinical Research (IZKF) of the medical faculty of Münster (grant Dan3/022/22 to UD).

The BiDirect Study was funded by grants from the German Federal Ministry of Education and Research (BMBF; grants FKZ-01ER0816, FKZ-01ER1205, and FKZ-01ER1506) to K.B.

This work was funded in part by the consortia grants from the German Research Foundation (DFG) FOR 2107, SFB/TRR 393 (“Trajectories of Affective Disorders”, project grant no 521379614, plus DFG KI 588/23-1), the Germany’s Excellence Strategy (EXC 3066/1 “The Adaptive Mind”, Project No. 533717223), the DYNAMIC center, funded by the LOEWE program of the Hessian Ministry of Science and Arts (grant number: LOEWE1/16/519/03/09.001(0009)/98) and the Wellcome Trust funded DIALOG consortium (Understanding Disorganisation: A Language-Focused Global Initiative in Psychosis; Grant Number 314138/Z/24/Z).

## Notes

### Competing Interest Statement

The authors have declared no competing interest.

### Author Declarations

Ethics committee of the Medical Faculty of the University of Muenster, the Westphalian Chamber of Physicians in Muenster, and Ethics committee of the Faculty of Medicine at the University of Marburg gave ethical approval for this work.

