## Supplemental Information for "The timeline of brain aging in major depression: A prospective study from before first-onset to established illness"

### SUPPLEMENTS

**Supplementary Table S1:** Demographic and clinical characteristics across the datasets at Baseline

| | HC | | MDD | | CON | |
|---|---|---|---|---|---|---|
| **Discovery (n=1528)** | n | mean (±SD) | n | mean (±SD) | n | mean (±SD) |
| Total | 903 | | 576 | | 49 | |
| Age | | 36.17 (13.05) | | 37.88 (13.41) | | 34.18 (11.13) |
| Female Sex | 539 | | 357 | | 27 | |
| HAM-D | | 1.17 (1.72) | | 9.84 (7.53) | | 1.80 (2.46) |
| BDI | | 3.46 (3.91) | | 18.55 (11.56) | | 5.29 (5.51) |
| **Replication (n=1190)** | n | mean (±SD) | n | mean (±SD) | n | mean (±SD) |
| Total | 514 | | 645 | | 31 | |
| Age | | 52.80 (8.69) | | 49.48 (7.55) | | 50.75 (8.10) |
| Female Sex | 250 | | 391 | | 19 | |
| CES-D | | 7.50 (5.96) | | 26.69 (12.22) | | 11.97 (7.25) |

**Supplementary Table S2:** Breakdown of scan exclusions across datasets.

| Exclusion Reason | Discovery Dataset | Replication Dataset |
|---|---|---|
| Missing age | 0 | 2 |
| Missing sex | 146 | 0 |
| brain age prediction failed | 82 | 77 |

**Supplementary Table S3:** Vulnerability - ANCOVA Results for Brain Age Gap at Baseline

| | df | F-value | p-value | Partial η2 |
|---|---|---|---|---|
| **Discovery Dataset** | | | | |
| Intercept | 1 | 13.48 | < .001 | — |
| Group | 2 | 5.26 | .006 | 0.02 |
| Chronological Age | 2 | 480.81 | < .001 | 0.65 |
| Biological Sex | 1 | 16.95 | < .001 | 0.03 |
| Total Intracranial Volume | 1 | 11.22 | .001 | 0.02 |
| Scanner Site | 3 | 20.26 | < .001 | 0.11 |

| | | | | |
|---|---|---|---|---|
| **Replication Dataset** | | | | |
| Intercept | 1 | 4.38 | .037 | — |
| Group | 2 | 4.49 | .012 | 0.03 |
| Chronological Age | 2 | 13.77 | < .001 | 0.08 |
| Biological Sex | 1 | 54.92 | < .001 | 0.14 |
| Total Intracranial Volume | 1 | 1.23 | .268 | < 0.01 |

**Supplementary Table S4:** Vulnerability - Post-Hoc Pairwise Marginal Means Comparisons Across Groups at Baseline

| | **Unstandardized Estimate (Years)** | **SE** | **df** | **t-ratio** | $p_{Tukey}$ | **Cohen's d** |
|---|---|---|---|---|---|---|
| **Discovery dataset** | | | | | | |
| HC vs. MDD | -1.39 | 0.44 | 507 | -3.14 | .005 | -0.29 |
| HC vs. CON | -0.03 | 0.76 | 507 | -0.04 | .999 | -0.01 |
| MDD vs. CON | 1.36 | 0.77 | 507 | 1.76 | .183 | 0.29 |
| **Replication dataset** | | | | | | |
| HC vs. MDD | -2.51 | 0.88 | 334 | -2.86 | .012 | -0.33 |
| HC vs. CON | 0.03 | 1.52 | 334 | 0.02 | .999 | 0.00 |
| MDD vs. CON | 2.54 | 1.52 | 334 | 1.68 | .217 | 0.33 |

**Supplementary Table S5:** Acute Consequence - Linear Mixed-Effects Model Results

| | **F** | $df_{num}$ | $df_{den}$ | **p-value** | **Partial η2** |
|---|---|---|---|---|---|
| **Discovery dataset** | | | | | |
| Intercept | 68.87 | 1 | 3.91 | .001 | — |
| Group | 4.16 | 2 | 532.42 | .016 | 0.02 |
| Time | 0.40 | 1 | 538.69 | .526 | < 0.01 |
| Chronological Age | 458.09 | 2 | 707.45 | < .001 | 0.56 |
| Biological Sex | 17.26 | 1 | 518.13 | < .001 | 0.03 |
| Total Intracranial Volume | 9.56 | 1 | 756.71 | .002 | 0.01 |
| Group x Time Interaction | 0.53 | 2 | 388.19 | .588 | < 0.01 |
| **Replication dataset** | | | | | |
| Intercept | 555.03 | 1 | 348.63 | < .001 | — |
| Group | 4.50 | 2 | 350.30 | .012 | 0.03 |
| Time | 0.67 | 1 | 419.87 | .413 | < 0.01 |
| Chronological Age | 16.38 | 2 | 442.43 | < .001 | 0.07 |

| | | | | | |
|---|---|---|---|---|---|
| Biological Sex | 56.46 | 1 | 338.03 | < .001 | 0.14 |
| Total Intracranial Volume | 5.17 | 1 | 429.61 | .024 | 0.01 |
| Group x Time Interaction | 0.33 | 2 | 249.21 | .720 | < 0.01 |

**Supplementary Table S6:** Acute Consequence - Estimated Longitudinal Slopes and Pairwise Comparisons Across Groups

| | **Slope Estimate (Δ Years/Year)** | **SE** | **df** | **t** | $p_{Tukey}$ | **Cohen's d (Slopes)** |
|---|---|---|---|---|---|---|
| **Discovery dataset** | | | | | | |
| ***Within-Group Slopes*** | | | | | | |
| Healthy Controls (HC) | 0.028 | 0.045 | 539 | — | — | — |
| Major Depressive Disorder (MDD) | -0.035 | 0.054 | 487 | — | — | — |
| Converters (CON) | -0.025 | 0.114 | 408 | — | — | — |
| ***Pairwise Slope Contrasts*** | | | | | | |
| HC vs. MDD | 0.063 | 0.063 | 390 | 1.00 | .575 | 0.05 |
| HC vs. CON | 0.053 | 0.118 | 387 | 0.45 | .894 | 0.04 |
| MDD vs. CON | -0.010 | 0.121 | 386 | -0.08 | .996 | -0.01 |
| **Replication dataset** | | | | | | |
| ***Within-Group Slopes*** | | | | | | |
| Healthy Controls (HC) | 0.082 | 0.100 | 420 | — | — | — |
| Major Depressive Disorder (MDD) | 0.179 | 0.104 | 412 | — | — | — |
| Pre-clinical Converters (CON) | 0.107 | 0.179 | 296 | — | — | — |
| ***Pairwise Slope Contrasts*** | | | | | | |
| HC vs. MDD | -0.097 | 0.121 | 249 | -0.80 | .702 | -0.05 |
| HC vs. CON | -0.025 | 0.190 | 249 | -0.13 | .990 | -0.01 |
| MDD vs. CON | 0.072 | 0.191 | 249 | 0.37 | .926 | 0.03 |

**Supplementary Table S7:** Acute Consequence - Cross-Sectional Associations Between Depression Severity Scales and Brain Age Gap at Follow-Up

| | df | F | p | Partial η2 | Partial r |
|---|---|---|---|---|---|
| **Discovery dataset** | | | | | |
| **HAM-D** | | | | | |
| HAM-D Score | 1 | 0.004 | .950 | < 0.01 | -0.0044 |
| Chronological Age | 2 | 125.368 | < .001 | 0.55 | — |
| Biological Sex | 1 | 7.217 | .008 | 0.03 | — |
| Total Intracranial Volume | 1 | 1.584 | .210 | 0.01 | — |
| Scanner Site | 2 | 12.146 | < .001 | 0.11 | — |
| **BDI** | | | | | |
| BDI Score | 1 | 0.095 | .759 | < 0.01 | 0.0223 |
| Chronological Age | 2 | 121.285 | < .001 | 0.56 | — |
| Biological Sex | 1 | 4.554 | .034 | 0.02 | — |
| Total Intracranial Volume | 1 | 2.567 | .111 | 0.01 | — |
| Scanner Site | 2 | 12.736 | < .001 | 0.12 | — |
| **Replication dataset** | | | | | |
| **CES-D** | | | | | |
| CES-D Score | 1 | 0.888 | .348 | 0.01 | -0.0872 |
| Chronological Age | 2 | 8.709 | < .001 | 0.13 | — |
| Biological Sex | 1 | 28.418 | < .001 | 0.20 | — |
| Total Intracranial Volume | 1 | 0.000 | .999 | < 0.01 | — |

**Supplementary Table S8:** Acute Consequence - Longitudinal Associations Between Changes in Depression Severity and Changes in Brain Age Gap

| | df | F | p | Partial η2 | Partial r |
|---|---|---|---|---|---|
| **Discovery dataset** | | | | | |
| **HAM-D** | | | | | |
| ΔHAM-D Score | 1 | 0.00 | .973 | < 0.01 | 0.00 |
| Baseline Age | 2 | 13.56 | < .001 | 0.12 | — |
| Biological Sex | 1 | 1.37 | .243 | 0.01 | — |
| Total Intracranial Volume | 1 | 2.87 | .092 | 0.01 | — |
| Scanner Site | 3 | 2.25 | .084 | 0.03 | — |
| **BDI** | | | | | |
| Δ BDI Score | 1 | 0.05 | .822 | < 0.01 | 0.02 |
| Baseline Age | 2 | 12.13 | < .001 | 0.12 | — |
| Biological Sex | 1 | 1.76 | .186 | 0.01 | — |
| Total Intracranial Volume | 1 | 1.38 | .242 | 0.01 | — |
| Scanner Site | 3 | 2.35 | .073 | 0.04 | — |
| **Replication dataset** | | | | | |
| **CES-D** | | | | | |
| ΔCES-D Score | 1 | 3.07 | .083 | 0.03 | 0.16 |
| Baseline Age | 2 | 1.82 | .167 | 0.03 | — |
| Biological Sex | 1 | 2.12 | .148 | 0.02 | — |
| Total Intracranial Volume | 1 | 0.03 | .860 | < 0.01 | — |

**Supplementary Table S9:** Residual - Mixed-Effects Model Results for Cumulative Depression Duration

| Fixed Effect Predictor | Estimate (β) | SE | df | t | p | Partial η2 |
|---|---|---|---|---|---|---|
| Intercept | -6.457 | 1.054 | 3.67 | -6.13 | .005 | — |
| Duration of Depressive Episodes | -0.005 | 0.003 | 270.80 | -1.54 | .126 | 0.01 |
| Chronological Age (Linear) | -132.700 | 6.383 | 393.60 | -20.80 | < .001 | 0.55 |
| Chronological Age (Quadratic) | 34.850 | 4.414 | 436.40 | 7.90 | < .001 | |
| Biological Sex (Male) | 2.080 | 0.634 | 279.10 | 3.28 | .001 | 0.04 |
| Total Intracranial Volume | 0.387 | 0.210 | 434.60 | 1.84 | .066 | 0.01 |

**Supplementary Table S10:** Residual - Mixed-Effects Model Results for Episode Recurrence

| Fixed Effect Predictor | Estimate (β) | SE | df | t | p | Partial η2 |
|---|---:|---:|---:|---:|---:|---:|
| Intercept *(No Relapse Ref.)* | -6.567 | 1.078 | 3.96 | -6.09 | .004 | — |
| Recurrence Status (Relapse) | 0.009 | 0.319 | 302.90 | 0.03 | .976 | < 0.01 |
| Recurrence Status (Healthy) | -0.085 | 0.348 | 244.50 | -0.24 | .808 | |
| Chronological Age (Linear) | -133.000 | 6.460 | 375.30 | -20.59 | < .001 | 0.55 |
| Chronological Age (Quadratic) | 34.500 | 4.452 | 436.50 | 7.75 | < .001 | |
| Biological Sex (Male) | 2.069 | 0.632 | 279.40 | 3.27 | .001 | 0.04 |
| Total Intracranial Volume | 0.399 | 0.213 | 436.40 | 1.88 | .062 | 0.01 |

**Supplementary Table S11:** Residual - Post-Hoc Pairwise Marginal Means Comparisons Across Groups for Episode Recurrence

| Contrast Comparison Groupings | Estimate (Δ) | SE | df | t | $p_{Tukey}$ |
|---|---:|---:|---:|---:|---:|
| No Relapse - Relapse | -0.009 | 0.320 | 303 | -0.03 | .999 |
| No Relapse - Healthy | 0.085 | 0.348 | 245 | 0.24 | .968 |
| Relapse - Healthy | 0.094 | 0.434 | 318 | 0.22 | .975 |

#### Supplementary Methods: Computation of Medication Load Index

To measure the total medication load, each psychotropic medication was coded as absent (0), low (1, representing an equal or lower average dose), or high (2, representing a greater than average dose) relative to the midpoint of the daily dose range recommended by the Physician's Desk Reference. A composite measure was calculated by summing these individual medication scores to reflect both the dose and variety of medications taken. The mean of the total medication scores was calculated across timepoints and multiplied by the percentage of days the participant received medication during the follow-up period.

#### Supplementary Methods: Detailed MRI Acquisition Parameters

In the MACS cohort, T1-weighted high-resolution anatomical data were acquired using three-dimensional fast gradient echo (MPRAGE) sequences on 3T scanners (Tim Trio, Siemens, Erlangen, Germany in Marburg; Prisma, Siemens, Erlangen, Germany in Münster). The sequence parameters for Marburg were: 176 sagittal slices, 0.5 mm slice gap, TR = 1900 ms, TE = 2.26 ms, inversion time = 900 ms, flip angle = 9°, and a voxel size of 1 x 1 x 1 $mm^3$. In Münster, the parameters were: 192 sagittal slices, 0.5 mm slice gap, TR = 2130 ms, TE = 2.28 ms, inversion time = 900 ms, flip angle = 8°, and a voxel size of 1 x 1 x 1 $mm^3$. In the MNC cohort, T1-weighted high-resolution anatomical images were acquired on a 3T scanner (Gyroscan Intera, Philips Medical Systems, the Netherlands) using a 3D turbo field echo (3D TFE) sequence. The parameters were: TR = 7.4 ms, TE = 3.4 ms, flip angle = 9°, two signal averages, an inversion prepulse every 814.5 ms, and reconstruction to voxels of 0.5 x 0.5 x 0.5 mm. In the BiDirect cohort, T1-weighted high-resolution anatomical images were acquired on the same 3T scanner as MNC. The parameters were: TR = 7.26 ms, TE = 3.56 ms, TI = 404ms flip angle = 9°, and reconstruction to voxels of 1 x 1 x 1 $mm^3$.

#### Supplementary Result: Age-Sex-Site Matching Effectiveness

Prior to matching, both datasets exhibited imbalances in chronological age (MACS+MNC: Kruskal-Wallis $chi^2$ = 6.42, p = .040; BiDirect: Kruskal-Wallis $chi^2$ = 43.08, p < .001), biological sex (MACS+MNC: Pearson $chi^2$ = 1.91, p = .385; BiDirect: Pearson $chi^2$ = 13.63, p = .001) and scanner site distribution (MACS+MNC: Pearson's $chi^2$ = 37.70, p < .001) at baseline. The matching procedure effectively neutralized these confounding variables.

Post-matching analyses confirmed that group differences in age were eliminated (MACS+MNC: Kruskal-Wallis $chi^2$ = 0.70, p = .70; BiDirect: Kruskal-Wallis $chi^2$ = 0.06, p = .970). Furthermore, distribution of biological sex (MACS+MNC: Pearson $chi^2$ = 0.02, p = .992; BiDirect: Pearson $chi^2$ = 0.132, p = .936) and across scanner sites (MACS+MNC: Pearson $chi^2$ = 0.38, p = .984) was thoroughly harmonized (see Supplementary Figure 1).

#### Supplementary Result: Pre-Matching Depressive Scores at Baseline

Pre-Matching clinical characterization confirmed significant differences in depressive symptomology across groups at baseline in the Discovery dataset. As expected, the chronic MDD group dataset exhibited moderate-to-severe symptom levels (BDI = 18.6 ± 11.6; HAMD = 9.8 ± 7.5). Notably, while the Converter group was recruited as healthy and remained well below clinical thresholds at baseline, they exhibited higher sub-clinical scores (BDI = 5.3 ± 5.5; HAMD = 1.8 ± 2.5) compared to the stable Healthy Control group (BDI = 3.5 ± 3.9; HAMD = 1.2 ± 1.7). In the Replication dataset the self-reported symptom severity showed even clearer differences between the groups (CES-D: HC = 7.5 ± 6.0; CON = 12.0 ± 7.3; MDD = 26.7 ± 12.2). Post-hoc comparisons showed the baseline disparities between healthy controls and pre-conversion subjects were statistically significant for self-reported scales in the Replication dataset (CES-D: $p_{Tukey}$ = .005) and displayed a marginal trend (BDI: $p_{Tukey}$ = .056) in the Discovery dateset, identifying a subtle but measurable symptomatic divergence prior to the first depressive episode.

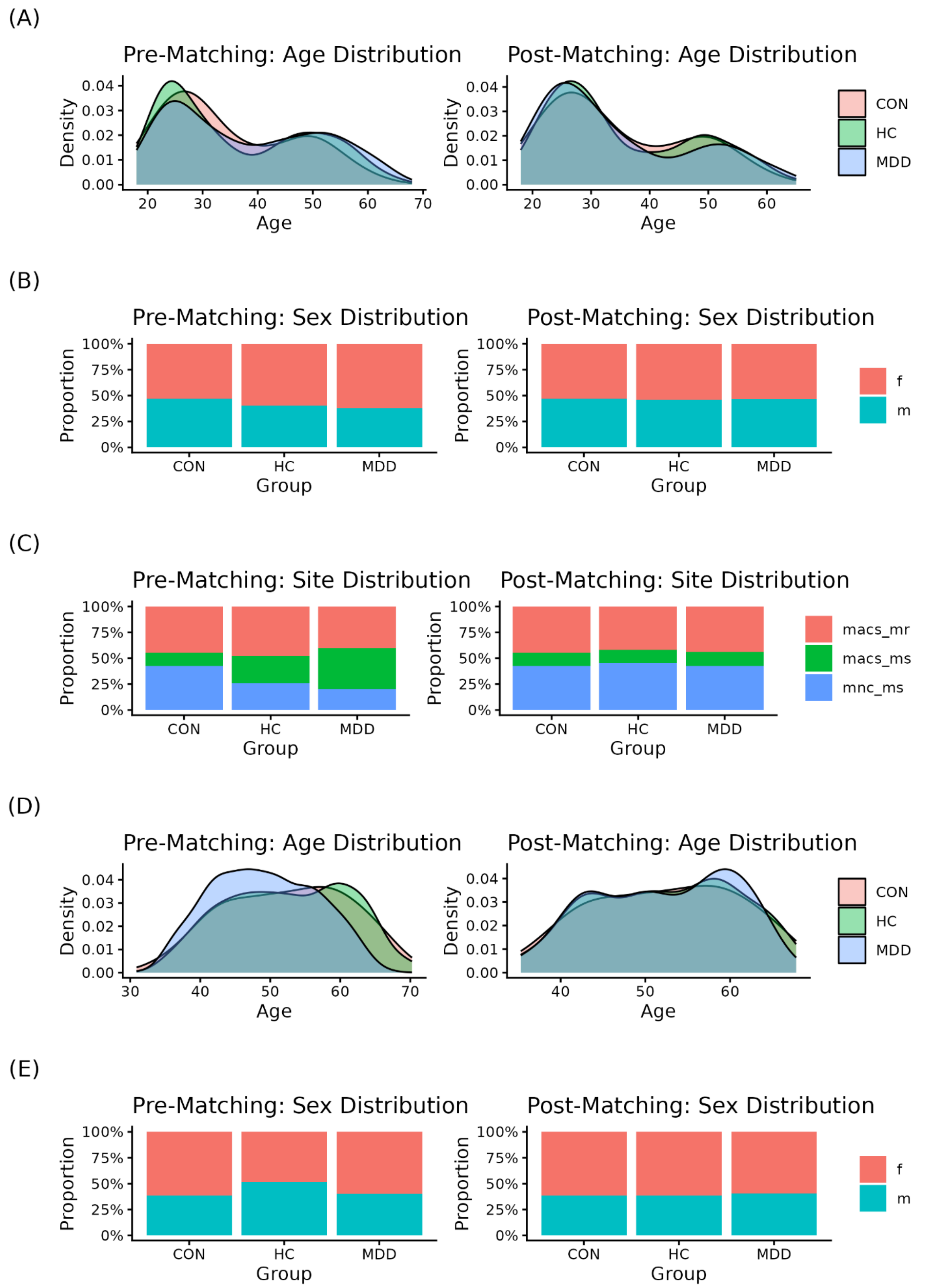

Supplementary Figure 1: Baseline demographic and technical distributions pre- and post-matching. Comparison of participant characteristics across the MACS/MNC (Panels A–C) and BiDirect (Panels D–E) cohorts. Distributions are shown for healthy controls (HC), patients with established major depressive disorder (MDD), and individuals who converted to

MDD during follow-up (CON). (A, D) Density plots of chronological age (years). (B, E) Proportional sex distribution (f: female, m: male). (C) Distribution of scanner site within the MACS/MNC cohort (macs_mr: MACS site, Marburg; macs_ms: MACS site, Münster; mnc_ms: MNC site, Münster). For each panel, the original baseline distribution is compared against the final matched sample to illustrate the achievement of group balance across demographic and technical covariates.
